# Efficient Prediction of Vitamin B Deficiencies via Machine-learning Using Routine Blood Test Results in Patients With Intense Psychiatric Episode

**DOI:** 10.1101/19004317

**Authors:** Hidetaka Tamune, Jumpei Ukita, Yu Hamamoto, Hiroko Tanaka, Kenji Narushima, Naoki Yamamoto

## Abstract

**Background:** Vitamin B deficiency is common worldwide and may lead to psychiatric symptoms; however, vitamin B deficiency epidemiology in patients with intense psychiatric episode has rarely been examined. Moreover, vitamin deficiency testing is costly and time-consuming, which has hampered effectively ruling out vitamin deficiency-induced intense psychiatric symptoms. In this study, we aimed to clarify the epidemiology of these deficiencies and efficiently predict them using machine-learning models from patient characteristics and routine blood test results that can be obtained within one hour.

**Methods:** We reviewed 497 consecutive patients deemed to be at imminent risk of seriously harming themselves or others over 2 years in a single psychiatric tertiary-care center. Machine-learning models (k-nearest neighbors, logistic regression, support vector machine, and random forest) were trained to predict each deficiency from age, sex, and 29 routine blood test results gathered in the period from September 2015 to December 2016. The models were validated using a dataset collected from January 2017 through August 2017.

**Results:** We found that 112 (22.5%), 80 (16.1%), and 72 (14.5%) patients had vitamin B_1_, vitamin B_12_, and folate (vitamin B_9_) deficiency, respectively. Further, the machine-learning models were well generalized to predict deficiency in the future unseen data, especially using random forest; areas under the receiver operating characteristic curves for the validation dataset (i.e. the dataset not used for training the models) were 0.716, 0.599, and 0.796, respectively. The Gini importance of these vitamins provided further evidence of a relationship between these vitamins and the complete blood count, while also indicating a hitherto rarely considered, potential association between these vitamins and alkaline phosphatase (ALP) or thyroid stimulating hormone (TSH).

**Discussion:** This study demonstrates that machine-learning can efficiently predict some vitamin deficiencies in patients with active psychiatric symptoms, based on the largest cohort to date with intense psychiatric episode. The prediction method may expedite risk stratification and clinical decision-making regarding whether replacement therapy should be prescribed. Further research includes validating its external generalizability in other clinical situations and clarify whether interventions based on this method could improve patient care and cost-effectiveness.

## 1. Introduction

Vitamin B deficiency is common worldwide and may lead to psychiatric symptoms^1–4^. For example, meta-analyses have shown that patients with schizophrenia or first-episode psychosis have lower folate (vitamin B_9_) levels than their healthy counterparts^4,5^. Moreover, vitamin therapy can effectively alleviate symptoms in a subgroup of patients with schizophrenia^3,6–8^. However, the epidemiology of vitamin B deficiency in patients with active mental symptoms requiring immediate hospitalization has rarely been examined.

In a psychiatric emergency, psychiatrists should promptly distinguish treatable patients with altered mental status due to a physical disease from patients with an authentic mental disorder (international statistical classification of diseases and related health problems-10, ICD-10 code: F2-9). However, vitamin deficiency testing is very costly (around 60 dollars for each measurement of vitamin B_1_ (vitB_1_), vitamin B_12_ (vitB_12_), or folate in the U.S.; 15–25 dollars for each test in Japan) and usually requires at least two days. Therefore, an efficient, cost-effective method of predicting vitamin B deficiency is needed.

Although several studies have applied machine-learning to the prediction of diagnosis or treatment outcomes^9–11^, no study using machine-learning has focused on vitamin B deficiencies. We herein explore whether vitB_1_, vitB_12_, and folate deficiencies can be predicted using a machine-learning classifier from patient characteristics and routine blood test results obtained within one hour based on a large cohort of patients requiring urgent psychiatric hospitalization.

## 2. Methods

### 2.1. Medical chart review

We reviewed consecutive patients admitted to the Department of Neuropsychiatry at Tokyo Metropolitan Tama Medical Center, one of the biggest psychiatric tertiary-care centers in Japan, between September 2015 and August 2017 under the urgent involuntary hospitalization law, which requires the immediate psychiatric hospitalization of patients at imminent risk of seriously harming themselves or others. The necessity of hospitalization was judged by designated mental health specialists. There were no exclusion criteria. The patient characteristics, ICD-10 codes, and laboratory data were gathered retrospectively.

Since the reference ranges for vitB_1_, vitB_12_, and folate are 70–180 nmol/L (30–77 ng/mL), 180–914 ng/L, and > 4.0 μg/L, respectively^12^, a deficiency of the nutrients was defined as < 30 ng/mL, < 180 ng/L, and < 4.0 μg/L, respectively, unless otherwise stated. The odds ratios of each deficiency in each ICD-10 code were calculated assuming binomial distribution.

### 2.2. Classifiers and statistics

We compared four types of standard machine-learning classifiers; k-nearest neighbors, logistic regression, support vector machine, and random forest. Each type of classifier was trained to predict the deficiency of each substance from age, sex, and 29 routine blood variables (described in the Result section with values). For developing the models, any missing values were replaced using the mean. The classifiers were trained using the dataset populated in the period from September 2015 to December 2016 (the “Training set”). First, except for logistic regression, we optimized the hyperparameters of the classifier by selecting the best combination of hyperparameters that maximized the “5-fold cross validation” accuracy, among many combinations within appropriate ranges. The cross-validation accuracy was computed as follows; in one session, the classifiers were trained using 80% of the training set and evaluated on the withheld 20% of the training set. This session was performed five times so that every data would be withheld once. The accuracies were finally averaged across sessions to yield the cross-validation accuracy. By incorporating this process, the classifiers were generalized to unseen data (Graphical method is shown in **Figure 1**).

**Figure 1:**
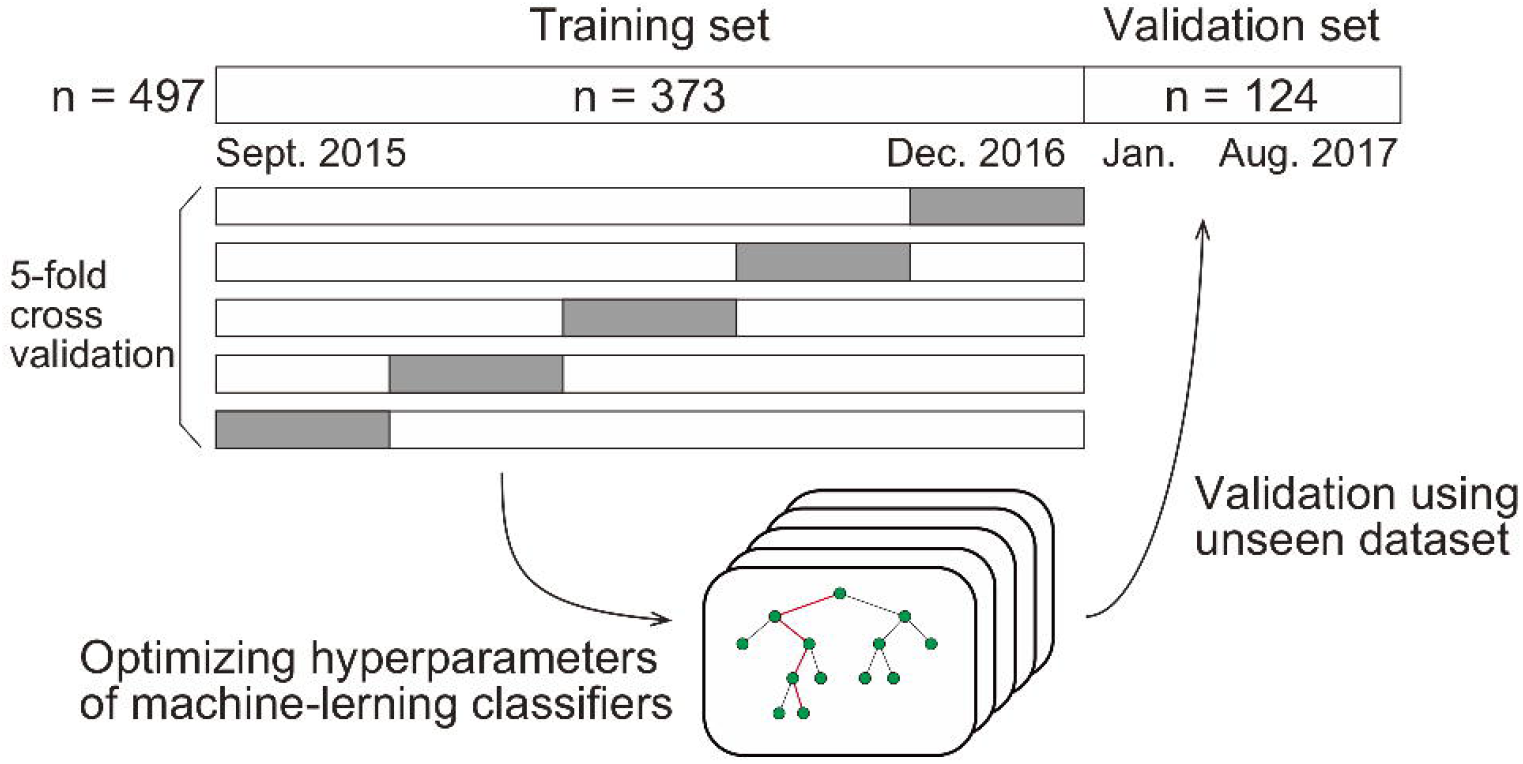
Graphical illustration of method of machine-learning.

Using the optimized hyperparameters, the classifiers were then validated using data collected from January 2017 through August 2017 (the “Validation set”). We report the classification performance on the validation set in the results section unless otherwise stated.

We quantified the sensitivity, specificity, and accuracy (defined as the average of the sensitivity and the specificity on the optimal operating point) using receiver operating characteristic curves (ROCs). We also quantified the 95% confidence interval of the area under the ROCs (AUCs) and accuracy using 1000-times bootstrapping.

When investigating the Gini importance and the partial dependency^13^, we retrained the classifiers using all datasets. All data analyses were performed using Python (2.7.10) with the Scikit-learn package (0.19.0) and R (3.4.2) with the edarf package (1.1.1) and pROC package (1.15.3).

### 2.3. Robustness verification

We verified the robustness of the prediction performances by three independent approaches. First, we compared the following two prediction performances; random forest classifiers trained and validated using the dataset from the F2 population, and random forest classifiers trained and validated using the dataset from the non-F2 population.

Second, we compared the prediction performances of several random forest classifiers trained and validated using the dataset where different cut-off values were used to define the vitamin deficiency. We chose other two cut-off values for each vitamin based on previous reports^14–16^, as well as pre-defined cut-off values (see also 2.1.).

Third, we trained and validated other random forest classifiers where the dataset was split in a different way. Here, the training set consisted of data between the 31^st^ of January, 2016 and August 2017 and the validation set consisted of data between September 2015 and the 31^st^ of January, 2016 so that the sample sizes of the training and validation sets were equal to those in the original split.

### 2.4. Subsampling analysis

We also examined the relationship between the dataset size and the generalization performance^17^. In this analysis, we trained the random forest classifiers using X% of the training set (X = 30, 35, 40, …, 95, and 100), and validated them using the validation set. The hyperparameters were identical to those used in the previous section. To remove sampling bias, this procedure was repeated 100 times for each value of X, where the training dataset was sampled randomly for each repetition. This results in obtaining 100 AUC scores for each X and for each vitamin. We plotted the AUC scores (averaged across the 100 repetition) versus X for each vitamin. Then the curve was fit with the following saturating function using Levenberg-Marquardt algorithm implemented as “curve_fit” function in the Scipy package (0.19.0).

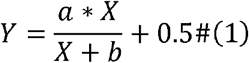

where *Y* is the AUC score, and *a* and *b* are the parameters to fit. Note that *Y* → *a* + 0.5 as *X* → ∞, and *Y* = 0.5 as *X* = 0.

### 2.5. Ethical considerations

Informed consent was obtained from participants using an opt-out form on the website. The study protocol was approved by the Research Ethics Committee, Tokyo Metropolitan Tama Medical Center (Approval number: 28-8). The study complied with the Declaration of Helsinki and the STROBE statement.

## 3. Results

### 3.1. Eligible patients

During the 2-year study period, 497 consecutive patients (496 were Asian) were enrolled. The mean age (standard deviation, SD) was 42.3 (±15.4) years, and 228 patients (45.9%) were women. F2 (Schizophrenia, schizotypal, delusional, and other non-mood psychotic disorders) was diagnosed in over 60% of the patients. The ICD-10 codes of the patients and the number of deficiencies at several cut-off values for vitB_1_, vitB_12_, and folate are shown in **Table 1**. According to the predefined cut-off values^12^, 112 (22.5%), 80 (16.1%), and 72 (14.5%) patients exhibited a deficiency of vitB_1_ (<30 ng/mL), vitB_12_ (<180 ng/L), and folate (<4.0 μg/L), respectively. Vitamin B deficiencies in sub-groups are shown in **Table 2**. A summary of the full dataset is shown in **Table 3**. Detailed information (sub-datasets) is shown in **Supplementary Table 1, 2, and 3** online. Histograms of vitB_1_, vitB_12_, and folate values are shown in **Figure 2 A-C**.

**Table 1.**
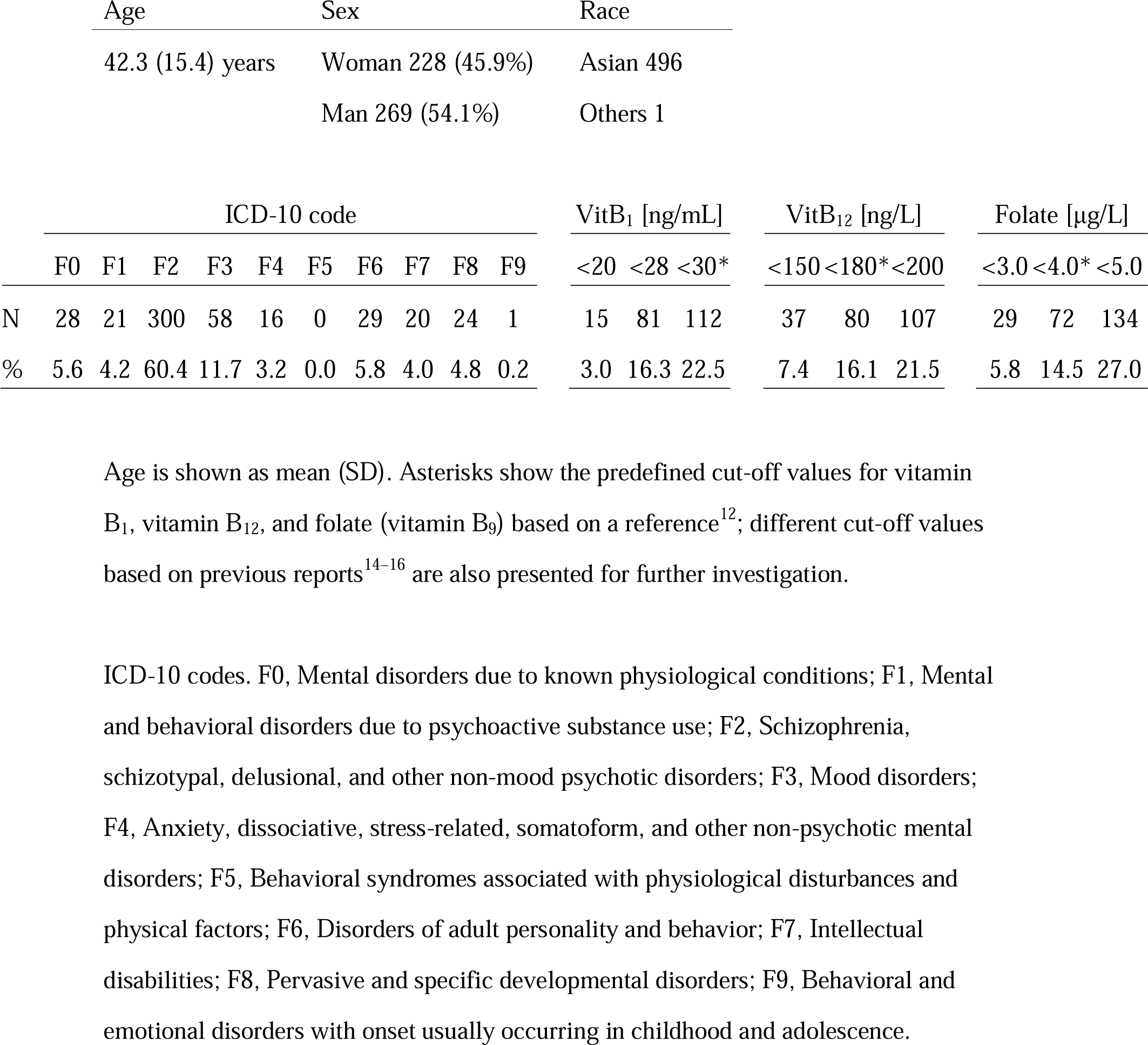
Patient distribution data (n = 497)

**Table 2.** Vitamin B deficiencies in sub-groups.

|  | F0 | F1 | F2 | F3 | F4 | F6 | F7 | F8 | F9 |
| --- | --- | --- | --- | --- | --- | --- | --- | --- | --- |
| vitB<sub>1</sub> < 30 | 9 | 4 | 70 | 11 | 3 | 7 | 5 | 3 | 0 |
| [ng/mL] | (32%) | (19%) | (23%) | (19%) | (19%) | (24%) | (25%) | (13%) |  |
| vitB<sub>12</sub> < 180 | 5 | 4 | 53 | 7 | 3 | 1 (3%) | 4 | 3 | 0 |
| [ng/L] | (18%) | (19%) | (18%) | (12%) | (19%) |  | (20%) | (13%) |  |
| Folate < 4.0 | 5 | 7 | 38 | 6 | 5 | 3 | 4 | 4 | 0 |
| [μg/L] | (18%) | (33%) | (13%) | (10%) | (31%) | (10%) | (20%) | (17%) |  |

| Odds ratio | F0 | F1 | F2 | F3 | F4 | F6 | F7 | F8 | F9 |
| --- | --- | --- | --- | --- | --- | --- | --- | --- | --- |
| vitB<sub>1</sub> < 30 | 1.68 | 0.80 | 1.12 | 0.78 | 0.79 | 1.10 | 1.15 | 0.48 | 0 |
| [ng/mL] | [0.74-3.83] | [0.26-2.43] | [0.73-1.73] | [0.39-1.57] | [0.22-2.81] | [0.46-2.65] | [0.41-3.24] | [0.14-1.63] |  |
| vitB<sub>12</sub> < 180 | 1.14 | 1.24 | 1.35 | 0.69 | 1.21 | 0.18 | 1.32 | 0.73 | 0 |
| [ng/L] | [0.42-3.10] | [0.41-2.43] | [0.82-2.23] | [0.30-1.58] | [0.34-4.35] | [0.02-1.31] | [0.43-4.05] | [0.21-2.52] |  |
| Folate < 4.0 | 1.30 | 3.16 | 0.70 | 0.65 | 2.81 | 0.59 | 1.32 | 1.04 | 0 |
| [μg/L] | [0.48-3.55] | [1.23-8.13] | [0.42-1.15] | [0.27-1.58] | [0.95-8.34] | [0.17-1.98] | [0.43-4.05] | [0.35-3.14] |  |

Square brackets indicate the 95% confidence interval.
Abbreviations; see **Table 1**.

**Table 3.** Summary of full dataset of age, sex, and 29 parameters for machine-learning.

| Parameters | Units | Mean | SD | UN | mg/dL | 12.9 | 6.7 |
| --- | --- | --- | --- | --- | --- | --- | --- |
| Age | years | 42.3 | 15.4 | Cre | mg/dL | 0.7 | 0.2 |
| Sex | Woman | n = 228 |  | T.bil | mg/dL | 0.7 | 0.4 |
| WBC | $\times 10^3/\mu\text{L}$ | 8.2 | 2.8 | Na | mmol/L | 139 | 3 |
| Hb | g/dL | 13.7 | 1.7 | Cl | mmol/L | 105 | 4 |
| Hct | % | 40.3 | 4.5 | K | mmol/L | 3.7 | 0.4 |
| MCV | fL | 89 | 6.6 | cor.Ca | mg/dL | 9.1 | 0.5 |
| Plt | $\times 10^4/\mu\text{L}$ | 24.9 | 6.3 | CK | IU/L | 514 | 1230 |
| RDW.CV | % | 13.5 | 1.3 | AST | IU/L | 31 | 34 |
| Neu | % | 70 | 11 | ALT | IU/L | 27 | 24 |
| Lym | % | 23 | 10 | LDH | IU/L | 239 | 91 |
| Mono | % | 6 | 2 | ALP | IU/L | 224 | 81 |
| Eo | % | 1 | 2 | γGTP | IU/L | 37 | 63 |
| Baso | % | 0 | 0 | Glu | mg/dL | 112 | 40 |
| TP | g/dL | 7.2 | 0.6 | CRP | mg/dL | 0.4 | 0.9 |
| Alb | g/dL | 4.4 | 0.4 | TSH | $\mu\text{IU/mL}$ | 1.7 | 2.4 |

Two patients lacked age data (no photo ID was available), and one patient lacked biochemistry data (inappropriate sample processing). For machine-learning, the missing values were replaced using the mean.
Abbreviations: WBC, white blood cell count; Hb, hemoglobin; Hct, hematocrit; MCV, mean corpuscular volume; RDW.CV, red blood cell distribution width-coefficient variation; Plt, platelet; Neu, neutrocyte fraction; Lym, lymphocyte fraction; Mono, monocyte fraction; Eo, eosinocyte fraction; Baso, basocyte fraction; TP, total protein; Alb, albumin; UN, urea nitrogen; Cre, creatinine; T.bil, total bilirubin; Na, sodium; Cl,
chloride; K, potassium; cor.Ca, corrected calcium; CK, creatine kinase; AST, aspartate transaminase; ALT, alanine transaminase; LDH, lactate dehydrogenase; ALP, alkaline phosphatase; γGTP, γ-glutamyltransferase; Glu, plasma glucose; CRP, C-reactive protein; TSH, thyroid-stimulating hormone.

**Figure 2:**
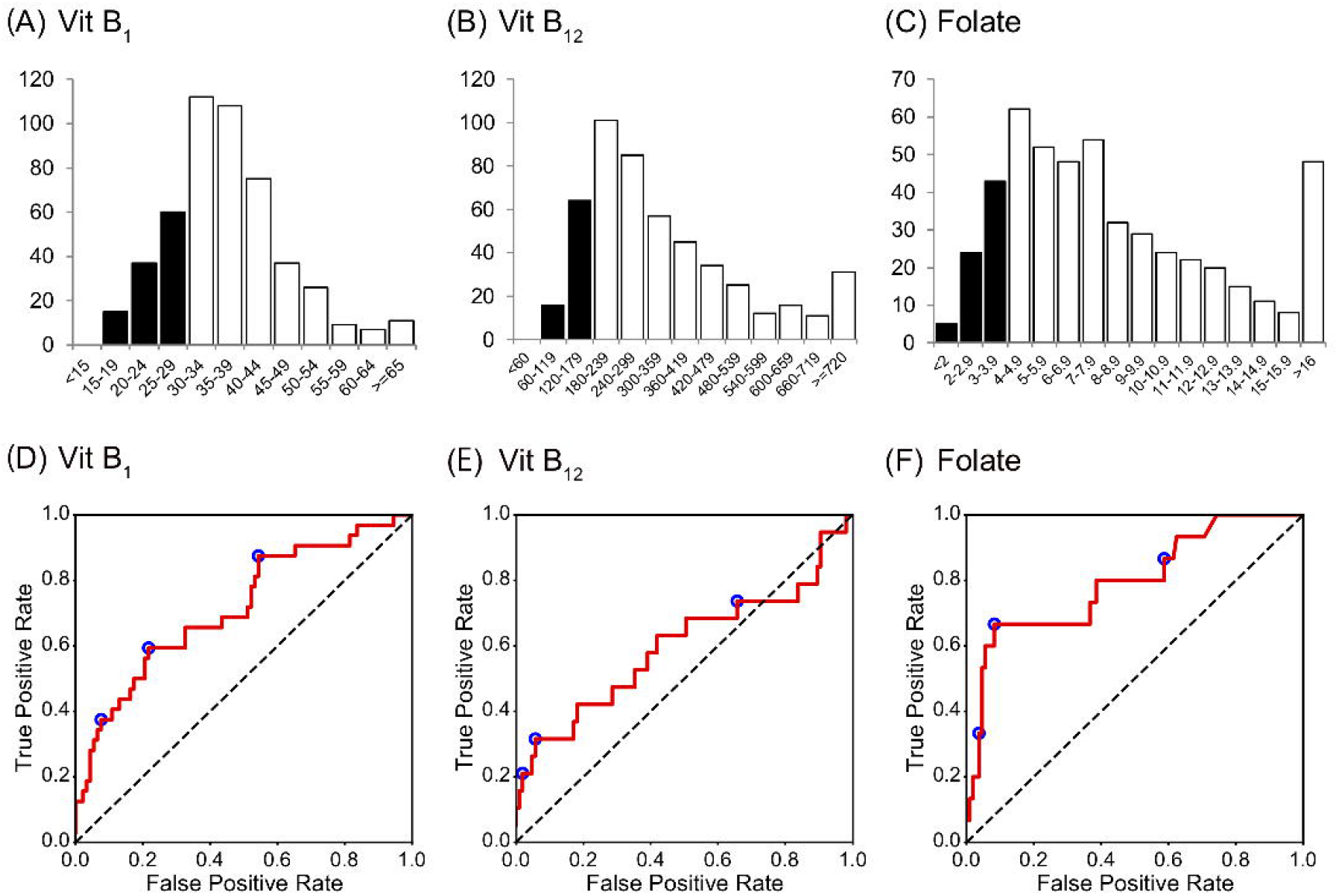
Histogram and ROC curves of each vitamin B value. (A-C) The histograms for vitamin B_1_, vitamin B_12_, and folate (vitamin B_9_). Their medians (1^st^–3^rd^ quartile) are 35 (30–42) ng/mL, 285 (206–431) ng/L, and 7.2 (4.9–10.8) μg/L, respectively. (D-F) ROC curves for vitamin B_1_, vitamin B_12_, and folate. Operating points used in **Table 4** and **Supplementary Table 5** are depicted in blue. Abbreviations: Vit B_1_, vitamin B_1_; Vit B_12_, vitamin B_12_.

### 3.2. Prediction via machine-learning using routine blood test results

Machine-learning classifiers were trained to predict the deficiency of each substance from patient characteristics and routine blood test results. The classifiers were trained using the dataset gathered in the period from September 2015 to December 2016 (the “Training set”, n = 373), which was then validated from January 2017 through August 2017 (the “Validation set”, n = 124). By splitting the whole dataset in this way, the ratio of the training and validation sample size was 3:1, a commonly used ratio in machine-learning analyses.

**Table 4.** Summary of AUC, sensitivity, specificity, and accuracy for the validation set.

| Classifier | vitB<sub>1</sub> | vitB<sub>12</sub> | Folate | Average |
| --- | --- | --- | --- | --- |
| k-nearest neighbors | 0.596<br>[0.483–0.702] | 0.542<br>[0.394–0.705] | 0.514<br>[0.383–0.651] | 0.551 |
| Logistic regression | 0.715<br>[0.602–0.815] | 0.602<br>[0.454–0.745] | 0.754<br>[0.610–0.877] | 0.690 |
| Support vector machine | 0.715<br>[0.613–0.814] | 0.620<br>[0.472–0.763] | 0.699<br>[0.536–0.842] | 0.678 |
| Random forest | 0.716<br>[0.610–0.825] | 0.599<br>[0.426–0.755] | 0.796<br>[0.656–0.911] | 0.704 |

Sensitivity, specificity, and accuracy for random forest
|  | vitB<sub>1</sub> | vitB<sub>12</sub> | Folate |
| --- | --- | --- | --- |
| Sensitivity | 0.594 | 0.316 | 0.667 |
| Specificity | 0.783 | 0.943 | 0.917 |
| Accuracy | 0.688<br>[0.597–0.787] | 0.629<br>[0.523–0.746] | 0.792<br>[0.665–0.909] |

Abbreviations: AUC, area under the receiver operating characteristic curve.

AUCs for the validation set for each classifier are summarized in **Table 4**. Although the performance of the classifiers was similar except for the k-nearest neighbors, random forest yielded the highest AUC on average. Therefore, we focused on random forest in the following analysis.

The AUCs of the random forest classifiers were 0.716, 0.599, and 0.796, for vitB_1_, vitB_12_, and folate, respectively (**Figure 2 D-F** and **Table 4**). With some operative points on the ROC, the sensitivity, specificity, and accuracy for the validation set were calculated (**Table 4**. See also **Supplementary Table 4** for training set and **Supplementary Table 5** for different operating points). The 95% confidence interval (CI) of the AUC and accuracy was quantified using 1000-times bootstrapping. For random forest classifiers, the 95% CI of each value did not include 0.5, except for the AUC of vitB_12_.

**Figure 3** shows the Gini importance (a–c) and partial dependency plots (d–f) for the eight most important variables for each substance. The results provided further evidence of a relationship between the vitamin B levels and complete blood count while also indicating the hitherto rarely considered, potential association between these vitamins and alkaline phosphatase (ALP) or thyroid stimulating hormone (TSH).

**Figure 3:**
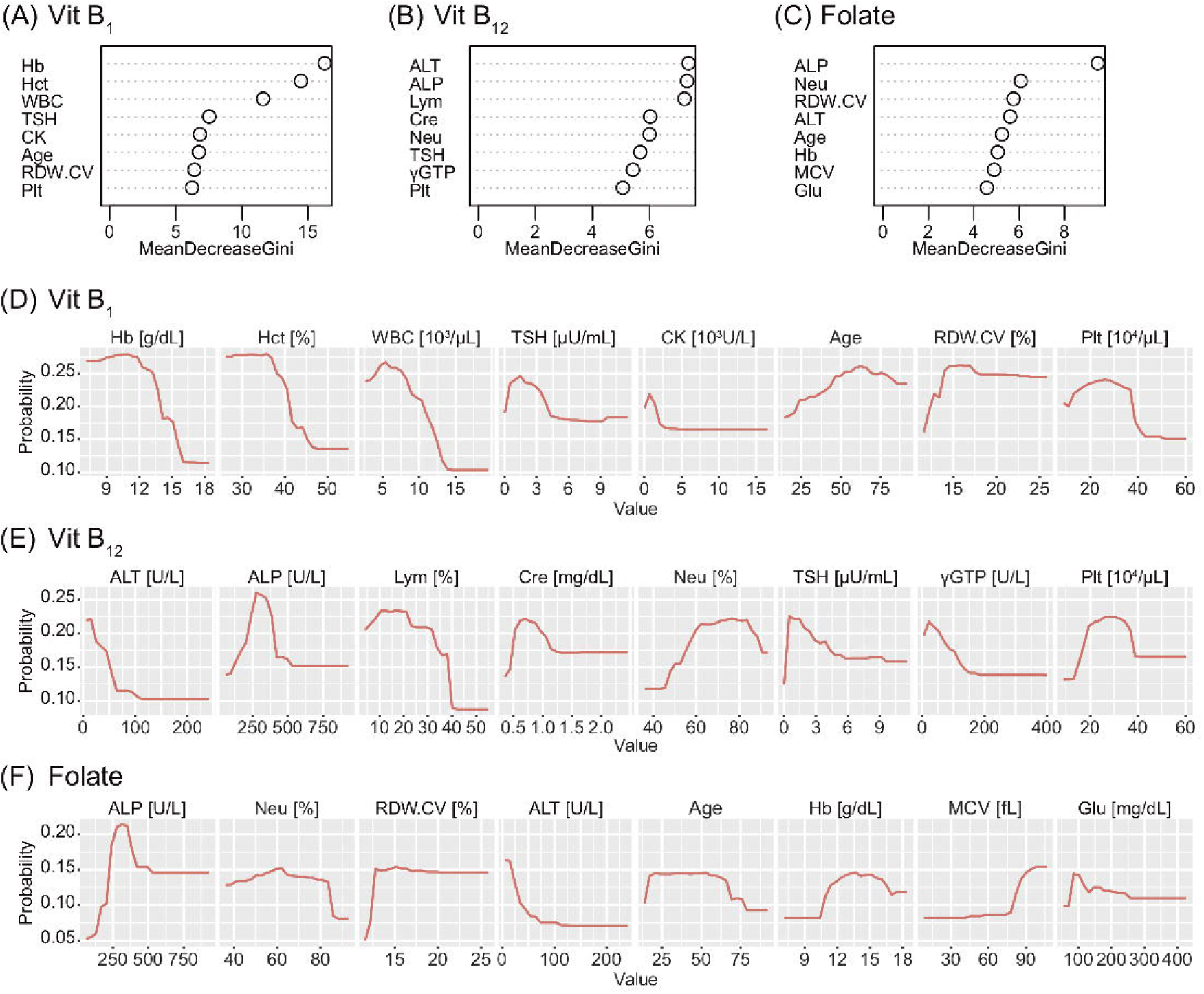
Gini importance and partial dependence plots of vitamin B deficiencies. The Gini importance (A-C) and partial dependency plots of the probability of deficiency (D-F) are shown for the eight most important variables for vitamin B_1_, vitamin B_12_, and folate (vitamin B_9_). Combined with these, this machine-learning classifier without hypothesis also provided further evidence of a relationship between vitamin B levels and the complete blood count while also indicating a potential association between these vitamins and alkaline phosphatase (ALP) or thyroid-stimulating hormone (TSH). Abbreviations: Vit B_1_, vitamin B_1_; Vit B_12_, vitamin B_12_; Hb, hemoglobin; Hct, hematocrit; WBC, white blood cell count; CK, creatine kinase; RDW.CV, red blood cell distribution width-coefficient variation; Plt, platelet; ALT, alanine transaminase; Lym, lymphocyte fraction; Cre, creatinine; Neu, neutrocyte fraction; γGTP, γ-glutamyltransferase; MCV, mean corpuscular volume; glu, plasma glucose.

### 3.3. Robustness verification

We verified the robustness of the results by three independent means. First, we asked if the prediction performance was influenced by the ICD-10 categories. When the prediction performances were compared between the random forest classifiers trained using the dataset from the F2 population and the classifiers trained using the dataset from the other population, the AUC was not statistically different (DeLong’s test), except in the case of vitB_1_ (see **Supplementary Table 6**).

Second, we used different cut-off values to define the deficiency^14–16^. Although the AUC for the validation set, shown in **Supplementary Table 7**, tended to be higher when strict cut-off values were used, the obtained AUCs were not statistically significant (p > 0.05, DeLong’s test with Bonferroni correction).

Third, we investigated if the prediction performance was influenced by the way the dataset was split into the training and validation set. Here, we trained and evaluated random forest classifiers using a dataset split in a reversed way (see Methods for details); The AUCs for the validation set were 0.771, 0.621, and 0.745 for vitB_1_, vitB_12_, and folate, respectively; none were statistically different from the AUC trained using the original setting (DeLong’s test), further demonstrating the robustness of the performance.

### 3.4. Subsampling analysis

To estimate the number needed to saturate the performance, we examined the relationship between the generalizability and the sample size^17^. We randomly sampled X% of the training set, trained random forest classifiers using the dataset, and evaluated the generalization performance by AUCs using the validation set (X = 30, 35, 40, …, 95, and 100; see Methods for details). As shown in **Figure 4**, the relationships between AUC and the training size for vitB_1_ and vitB_12_ were almost saturated, whereas that for folate is not saturated. To quantitatively understand this, we fitted each curve using a saturating function formulated in equation (1) (see Methods for details). The fitted parameters of equation (1) were as follows; for vitB_1_, a = 0.186 and b = 0.074; for vitB_12_, a = 0.099 and b = 0.156; and for folate, a = 0.291 and b = 0.123. By using these parameter values and extrapolating the curve, we then computed how many additional samples are necessary to reach almost maximum performances. To reach 99% of the maximum performance (i.e. Y = (a + 0.5) × 0.99 in equation (1)), the training dataset to be collected was 92.5%, 143%, and 341% of the training size in this study for vitB_1_, vitB_12_, and folate, respectively. These quantitative analyses revealed that collecting further similar datasets up to 1,000 patients (e.g. four years × hospitals with similar scale as Tokyo Metropolitan Tama Medical Center) may increase and reproduce the generalizability for folate, while the effect of collecting further dataset is expected to be small for vitB_1_ and vitB_12_.

**Figure 4:**
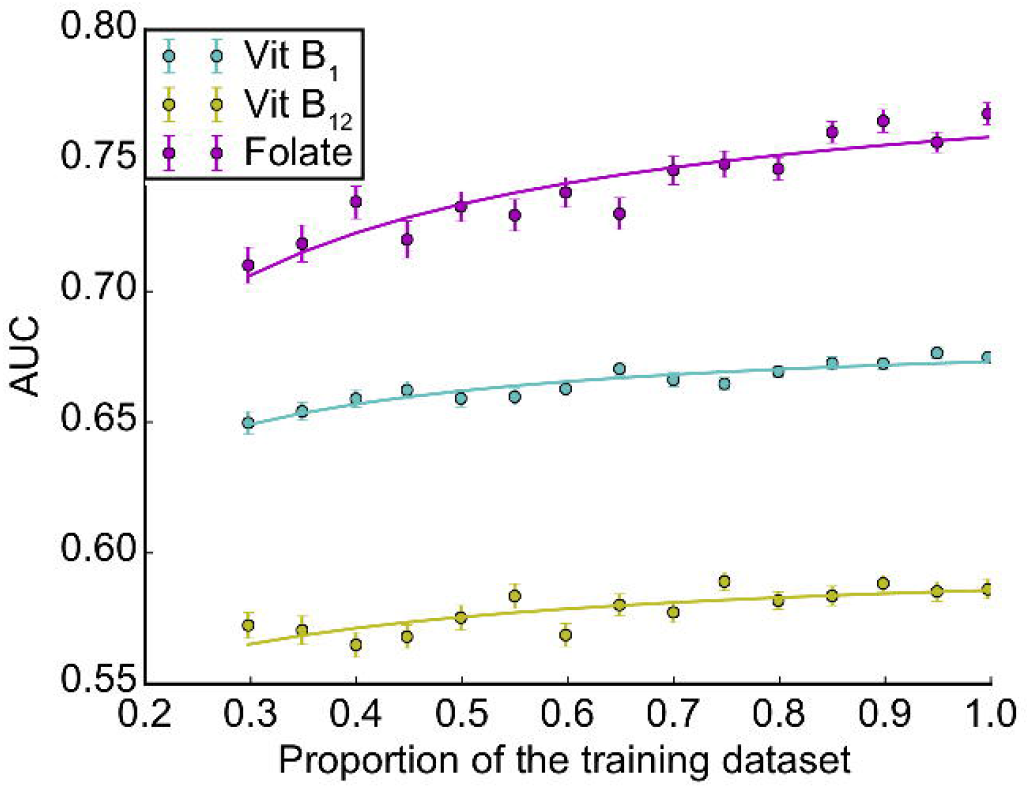
Subsampling analysis. The AUC performances as a function of the dataset size is shown for each vitamin (mean ± SEM across 100 repetitions; see Methods for details).

## 4. Discussion

### 4.1. Relevance of the present study

Based on the largest cohort to date of patients at imminent risk of seriously harming themselves or others, this study indicated that deficiency of certain vitamins can be predicted in an efficient manner via machine-learning using routine blood test results. The 29 routine blood variables are available at almost all hospitals/clinics and are necessary to rule out other comorbid physical problems. Given the large number of patients with vitamin B deficiencies, empirical therapy might be acceptable; however, risk stratification is preferred for personalized medicine and shared decision-making. The prediction method presented here may expedite clinical decision-making as to whether vitamins should be prescribed to a patient (Graphical abstract is shown in **Figure 5**).

**Figure 5:**
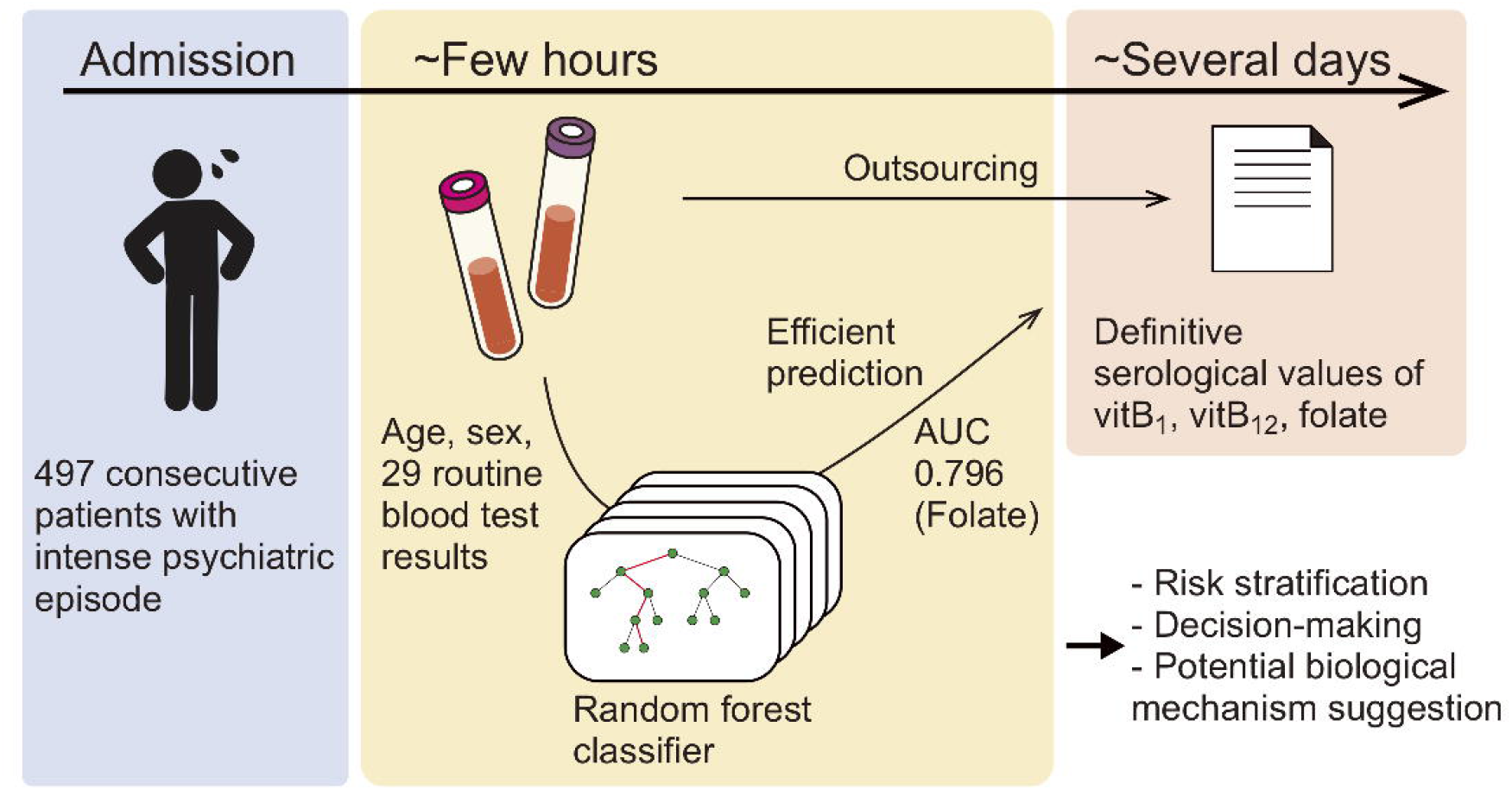
Graphical abstract.

Remarkably, the AUC of folate deficiency was 0.796. The robustness of folate prediction was also suggested by various independent methods and statistics. Folate has a potential to maintain neuronal integrity and is one of the homocysteine-reducing B-vitamins^5^. Homocysteine may be linked to the etiology of schizophrenia^18^, and vitamin B supplements have been reported to reduce psychiatric symptoms significantly in patients with schizophrenia^7^. A recent meta-review has pointed out that the bioactivity of the supplement should be considered (e.g. methylfolate, which successfully crosses the blood-brain barrier, has been reported effective, whereas the effect of other forms of folate is equivocal)^19^. As our study does not present longitudinal clinical courses, an intervention effect of folate supplementation to the cohort based on our method remains to be clarified.

### 4.2. Biological mechanism prediction

To connect with biological knowledge, we compared four models with high interpretability in this study. Using the random forest classifiers, as shown in **Figure 3**, we identified several items related to complete blood count as top hits. Notably, our classifier was blind to any biological knowledge, including the well-established association between anemia and vitamin B deficiency, including folate^20^. The results provide further evidence of a relationship between vitamin B levels and the complete blood count and support the use of machine-learning to investigate novel, underlying biological mechanisms^21^.

ALP and its metabolites indicate the vitamin B_6_ status^22^; low vitB_12_ is potentially associated with low ALP^23^. More generally, ALP may have a close and complicated relationship with the overall vitamin B group. Autoimmune disorders, especially thyroid disease, are commonly associated with pernicious anaemia^24^, but there has been no established hypothesis regarding the causal relationships between thyroid disease and vitamin B deficiencies. The potential association between the levels of these vitamins and ALP or TSH awaits further study, both via investigations of populations and basic research^25^.

### 4.3. Limitations

This study is subject to several limitations. First, the findings of this single-center retrospective study may have limited external generalizability, though internal generalizability was considered to the maximum extent. Second, the patients’ basic characteristics and long-term prognosis were not fully investigated due to administrative restrictions. Though there is similar involuntary treatment/admission in psychiatry worldwide, there is a gap between legislation and practice^26^. Therefore, the extent to which this method can expedite clinical decision-making is unclear.

Further, we did not investigate the relationship between serological values and the need for intervention. The lack of data for vitamin B deficiency in the Japanese general population hampered the comparison between the experimental cohort and their counterparts who lacked psychiatric symptoms. Establishing appropriate reference values and an assessment method requires further investigation. Finally, we did not assess the predictive value of other nutritional impairments, including vitamin B_6_ and homocysteine deficiency, which were previously shown to have a close link with psychiatric symptoms^3,5^; however, our study provides fundamental data on nutritional impairment based on the largest cohort of patients with intense psychiatric episode ever assembled for this purpose and presents a potential framework for predicting nutritional impairment using machine-learning.

### 4.4. Conclusion

The present report is, to the best of our knowledge, the first to demonstrate that machine-learning can efficiently predict nutritional impairment. This study also provided the possible application of machine-learning to investigate novel, underlying biological mechanisms. Further research is needed to validate the external generalizability of the findings in other clinical situations and clarify whether interventions based on this method can improve patient care and cost-effectiveness.

## Data Availability

The source code will be available on https://github.com/ukky17/vitaminPrediction after acceptance. The datasets utilized in the current study are available from the corresponding author upon reasonable request.

## 5. Contribution to the Field Statement

Vitamin B deficiency is common worldwide and may lead to psychiatric symptoms; however, vitamin B deficiency epidemiology in patients with intense psychiatric symptoms has rarely been examined. Moreover, vitamin deficiency testing is costly and time-consuming. Based on the largest cohort to date of patients at imminent risk of seriously harming themselves or others, this study demonstrated that the deficiency of certain vitamins can be predicted in an efficient manner via machine-learning models from patient characteristics and routine blood test results obtained within one hour.

In detail, among the 497 patients investigated (over 60% was diagnosed with schizophrenia or related psychotic disorders), 22.5%, 16.1%, and 14.5% patients had a deficiency of vitamin B_1_, B_12_, and folate, respectively, by direct measurement. Further, the machine-learning models well generalized to predict the deficiency in unseen datasets; areas under the receiver operating characteristic curves (AUCs) for the validation dataset were 0.716, 0.599, and 0.796, respectively. The prediction method presented in this study may expedite risk stratification and clinical decision-making regarding whether replacement therapy should be prescribed. The results also provided further evidence for well-known relationships between these vitamins and the complete blood count and supported the application of machine-learning to investigate novel, underlying biological mechanisms.

## 6. Acknowledgements

We thank Mr. James Robert Valera for his assistance in editing this manuscript and all the staff for their care of the patients and their contributions to this study. This manuscript has been previously published as a preprint^27^ in medRxiv (https://doi.org/10.1101/19004317).

## 7. Author Contributions Statement

H. Tamune has full access to all data and takes responsibility for the integrity of the data. H. Tamune, JU, KN, and NY conceived the study. H. Tamune, YH, and H. Tanaka collected the data. JU performed the statistical analyses. H. Tamune and JU drafted the first version of the manuscript. All authors critically revised the manuscript for intellectual content and approved the final version.

## Supporting Material List

Supplementary Table 1 (related to Table 1). Divided patient distribution data (n = 497)

Supplementary Table 2 (related to Table 2). Divided data of vitamin B deficiencies in sub-groups

Supplementary Table 3 (related to Table 3). Divided dataset of age, sex, and 29 parameters

Supplementary Table 4 (related to Table 4). Summary of sensitivity, specificity, and accuracy for the training set

Supplementary Table 5 (related to Table 4). Sensitivities and specificities at other operating points

Supplementary Table 6. Subgroup analyses

Supplementary Table 7. AUC with different cut-off values

